# The association of typical and atypical symptoms on in-hospital mortality of older adults with COVID-19: a multicentre cohort study

**DOI:** 10.1101/2022.07.16.22277716

**Authors:** Eric Kai-Chung Wong, Jennifer Watt, Hanyan Zou, Arthana Chandraraj, Alissa Wenyue Zhang, Jahnel Brookes, Ashley Verduyn, Anna Berall, Richard Norman, Katrina Piggott, Terumi Izukawa, Sharon E. Straus, Barbara Liu

**Affiliations:** Li Ka Shing Knowledge Institute, St. Michael’s Hospital, Unity Health Toronto, Toronto, Canada; Division of Geriatric Medicine, Department of Medicine, St. Michael’s Hospital, Unity Health Toronto, Toronto, Canada; Division of Geriatric Medicine, Department of Medicine, University of Toronto, Toronto, Canada; Division of Geriatric Medicine, Department of Medicine, Sinai Health and University Health Network, Toronto, Canada; Division of Geriatric Medicine, Department of Medicine, Sunnybrook Health Sciences Centre, Toronto, Canada; Kunin-Lunenfeld Centre for Applied Research & Evaluation, Rotman Research Institute, Baycrest Health Sciences Centre, Toronto, Canada; Providence Healthcare and Houses of Providence, Unity Health Toronto, Toronto, Canada; Division of Geriatric Medicine, Department of Medicine, Baycrest Health Sciences Centre, Toronto, Canada

**Author notes:** Corresponding author Barbara Liu, Professor, Department of Medicine, University of Toronto, Sunnybrook Health Sciences Centre, 2075 Bayview Ave., Room H4 79, Toronto, ON M4N 3M5.

**Keywords:** COVID-19, geriatrics, atypical presentations, delirium, geriatric syndromes

## Abstract

Atypical disease presentations are common in older adults with COVID-19. The objective of this study was to determine the prevalence of atypical and typical symptoms in older adults with COVID-19 through progressive pandemic waves and the association of these symptoms with in-hospital mortality. This retrospective cohort study included consecutive adults aged over 65 years with confirmed COVID-19 infection who were admitted to seven hospitals in Toronto, Canada from March 1, 2020 to June 30, 2021. The median age for the 1786 patients was 78.0 years and 847 (47.5%) were female. Atypical symptoms (as defined by geriatric syndromes) occurred in 1187 patients (66.5%), but rarely occurred in the absence of other symptoms (n=106, 6.2%). The most common atypical symptoms were anorexia (n=598, 33.5%), weakness (n=519, 23.9%), and delirium (n=449, 25.1%). Dyspnea (adjusted odds ratio [aOR] 2.05, 95% confidence interval [CI] 1.62–2.62), tachycardia (aOR 1.87, 95% CI 1.14–3.04), and delirium (aOR 1.52, 95% CI 1.18–1.96) were independently associated with in-hospital mortality. In a cohort of older adults hospitalized with COVID-19 infection, atypical presentations frequently overlapped with typical symptoms. Further research should be directed at understanding the cause and clinical significance of atypical presentations in older adults.

## Introduction

COVID-19 infection is associated with increased morbidity and mortality in older adults (Guan *et al*., 2020). Studies early in the pandemic revealed that older adults frequently did not have typical symptoms, such as cough, fever, or shortness of breath (Singhal *et al*., 2021). Atypical symptoms in older adults include geriatric syndromes, such as delirium, anorexia, or falls (Singhal *et al*., 2021). These presentations are non-specific to COVID-19 (Hofman *et al*., 2017) and can occur with other diagnoses; as such, clinicians needed to increase their level of suspicion of COVID-19 infection in older adults. Atypical presentations in COVID-19 have been variably associated with mortality (Gan *et al*., 2020). However, the definition of atypical presentation differs by study and it is often mixed with non-geriatric syndromes, such as abdominal pain or headache (Gan *et al*., 2020). A geriatric-focused approach to the classification of COVID-19 symptoms would help better illustrate the spectrum of presentations.

As recurrent waves of the pandemic occurred across the world, new variants of COVID-19 emerged. Later variants, particularly the beta (B.1.351), gamma (P.1), and delta (B.1617.2) variants, were more virulent (Fisman and Tuite, 2021) and presented with different symptoms (ZOE COVID Study, 2021). In Ontario, Canada, wave 1 was predominantly caused by wild-type SARS-CoV-2 virus, while wave 2 was predominantly alpha variant (B.1.1.7) and wave 3 had increasing beta and gamma variants (Health Ontario, 2021). It is unknown whether clinical presentation changed in older adults across these three waves.

Furthermore, despite systematic reviews of early studies of COVID-19 symptoms (Singhal *et al*., 2021), there is a lack of data looking at the way individuals with different comorbidities, place of residence, and frailty present. In particular, older age has been cited as the primary factor associated with atypical presentations of COVID-19, but it is unclear whether comorbidities and frailty also play a role.

Using a multicentre study of consecutively admitted older adults with COVID-19, the objectives of this study were to determine (i) the prevalence of each symptom and symptom category in older adults with COVID-19, (ii) the association of each symptom with in-hospital mortality, (iii) the change in symptoms with progressive waves of the pandemic and (iv) whether clinical presentation differs by age, frailty, place of residence and comorbidity.

## Methods

This retrospective cohort study was done across seven hospitals in Toronto, Canada from March 1, 2020 to June 30, 2021. Two sites had long-term care and rehabilitation beds (Providence Healthcare and Baycrest Health Sciences). The remaining hospitals were acute care institutions (St. Michael’s Hospital, Mount Sinai Hospital, Sunnybrook Hospital, Toronto General Hospital, and Toronto Western Hospital). Each of these facilities cared for patients with COVID-19 infections. Research ethics approval was obtained through Clinical Trials Ontario (3186-OPIA-Apr/2020-38044). The protocol for this study was published on Open Science Framework (https://osf.io/k4g7a/).

### Inclusion criteria

1. Patients with COVID-19 infection confirmed by viral polymerase chain reaction (PCR) swab available from hospital medical records. Patients received COVID-19 tests either by screening protocols or by clinical suspicion of disease.
2. Age ≥65 years at the time of COVID-19 detection.
3. Admitted to one of the acute care hospitals, rehabilitation facilities, or LTC homes listed above.

### Exclusion criteria

1. Re-admission to hospital after index admission for COVID-19. Only charts from the initial admission were included.

### Data collection

Decision support services at each hospital network identified COVID-19 cases in patients age 65 years or older on a monthly basis. A trained chart assessor conducted the chart review of each identified case and completed the case report forms hosted on a secure REDCap server at the Applied Health Research Centre in St. Michael’s Hospital.

Since reports in the literature used various definitions of typical and atypical symptoms (Davis *et al*., 2020; Pritchard *et al*., 2020), we created 3 categories of symptoms: (i) classic, (ii) typical, and (iii) atypical. Classic symptoms (sometimes referred to as “typical symptoms” in the literature) included fever, cough and shortness of breath. Our definition of typical symptoms included other symptoms (sometimes labelled as “atypical symptoms” in the literature) defined by the World Health Organization case definition of COVID-19 infection (e.g. rhinorrhea, diarrhea, nausea, and chills) (World Health Organization, 2020). We reserved the category of atypical symptoms for geriatric syndromes reported in the literature (Emmett, 1998; Hofman *et al*., 2017), such as delirium, anorexia, and weakness. Symptoms were extracted from each patient’s chart, along with clinical frailty scale (CFS) (Rockwood *et al*., 2005), place of residence and significant past medical history. We used CFS classification from the original study (Rockwood *et al*., 2005).

The waves of the pandemic were defined by dates provided by Toronto Public Health or on the date of the nadir of case counts between waves if a public health definition was not available. Wave 1 occurred from March 11, 2020 to July 31, 2020, using dates defined by Toronto Public Health (City of Toronto, 2021). Wave 2 spanned August 1, 2020 to February 20, 2021, which was defined by the nadir of case counts. Wave 3 occurred from February 21, 2021 until June 30, 2021, which was our predefined study end date.

The primary outcome was in-hospital mortality. Each chart was assessed for delirium on presentation from emergency room and admission notes using a validated chart review method (CHART-DEL) (Saczynski *et al*., 2014). Each chart assessor completed a training module for determination of CFS and extraction of CFS was regularly reviewed with a physician investigator. A training exercise was done on five charts where one of the physician investigators (BL, JW, EW, KP, TI, and AV) extracted chart data in duplicate with the chart assessor. After, the chart assessor regularly reviewed abstraction questions with the investigators. Five of the physician investigators were geriatricians (BL, JW, EW, KP, and TI) and one was a family physician (AV).

### Data processing

The dataset was reviewed for any missing variables. Missing key variables (e.g. CFS, delirium, death) were flagged and those charts were reviewed by a physician investigator. Error checking was done to identify variables outside of acceptable ranges. Summary variables were created for any atypical presentation, any typical presentation, any classic symptoms and any functional impairment. Missing CFS data was imputed as 6 (moderate frailty) for residents from LTC and 5 (mild frailty) for those from retirement homes or assisted living facilities based on local LTC admission criteria and published frailty estimates (Muscedere *et al*., 2016). Missing frailty or functional status data from community dwelling patients were not imputed because of the diverse range of frailty levels in this cohort (Kelly *et al*., 2017).

### Analysis

Patient baseline characteristics were analysed descriptively with proportions, means (standard deviations), and medians (interquartile range), where appropriate. Comparisons of categorical variables were done by chi-squared test and of continuous variables by ANOVA test. Non-normally distributed variables such as length of stay were compared using the Kruskal-Wallis test. The prevalence of each symptom was calculated by a proportion of exhibiting patients compared with the whole group.

Multivariable logistic regression models were used to identify independent association of symptoms for in-hospital mortality in older adults. Covariates of interest include age, sex, CFS, and total comorbidity count (based on documented past medical history). In the multivariable models, missing data were handled by listwise deletion. Statistical significance was defined at p ≤ 0.05. The analysis was done using R version 4.0.3.

Bubble plots were used to demonstrate the prevalence of typical and atypical symptoms in individuals grouped by age, frailty, place of residence and comorbidity. For this analysis, we combined classic and typical symptoms into a single “typical symptoms” category so that we could plot the findings on two axes. The sample size of each plotted group was reflected in the circle size. Bubble plots were generated by Microsoft Excel.

### Reporting standard

The Strengthening the Reporting of Observational Studies in Epidemiology (STROBE) Statement was used to write this manuscript (Von Elm *et al*., 2007).

## Results

### Baseline characteristics

A total of 1786 patients (Table 1) were included; their median age was 78.0 years (interquartile range [IQR] 71.0–85.0) and 847 (47.5%) were female. Overall, most patients were admitted from the community (n=1247, 70.0%) or LTC (n=258, 14.5%). The mean preadmission CFS score was 4.73 (standard deviation [SD] 1.65). The median number of comorbidities per patient was 2.0 (IQR 1.0–4.0).

**Table 1:** Prevalence of individual symptoms of COVID-19 presentation in hospitalized older adults stratified by in-hospital mortality. Place of residence indicates location of patient prior to admission.

| Symptom | Missing, n (%) | Cohort, n (%) | Death, n (%) | No death, n (%) | p-value |
| --- | --- | --- | --- | --- | --- |
| n |  | 1786 (100) | 418 (23.4) | 1359 (76.1) |  |
| Baseline characteristics |  |  |  |  |  |
| Age, median (IQR) | 0 | 78.0 (71.0–85.0) | 82.0 (74.0–89.0) | 76.0 (70.0–84.0) | <0.001 |
| Female sex*, n (%) | 0 | 847 (47.5) | 181 (43.3) | 663 (48.8) | 0.055 |
| Clinical frailty scale, mean (SD) | 61 (3.4) | 4.73 (1.65) | 5.28 (1.68) | 4.56 (1.60) | <0.001 |
| Number of comorbidities, median (IQR) | 0 | 2.0 (1.0–4.0) | 3.0 (2.0–4.0) | 2.0 (1.0–4.0) | <0.001 |
| Place of residence | 5 (0.3) |  |  |  | <0.001 |
| Community |  | 1247 (70.0) | 241 (58.1) | 997 (73.5) |  |
| LTC |  | 258 (14.5) | 88 (21.2) | 170 (12.5) |  |
| Hospital |  | 100 (5.6) | 28 (6.7) | 72 (5.3) |  |
| Retirement home |  | 96 (5.4) | 34 (8.2) | 62 (4.6) |  |
| Rehab |  | 47 (2.6) | 17 (4.1) | 30 (2.2) |  |
| Shelter |  | 22 (1.2) | 2 (0.5) | 20 (1.5) |  |
| Other institution |  | 10 (0.6) | 5 (1.2) | 5 (0.4) |  |
| Symptom classification |  |  |  |  |  |
| Any classic symptoms, n (%) | 0 | 1412 (79.1) | 348 (83.3) | 1057 (77.8) | 0.019 |
| Any typical symptoms, n (%) | 0 | 1263 (70.7) | 291 (69.6) | 965 (71.0) | 0.628 |
| Any atypical symptoms, n (%) | 0 | 1187 (66.5) | 295 (70.6) | 885 (65.1) | 0.045 |
| Classic symptoms |  |  |  |  |  |
| Cough | 0 | 929 (52.0) | 211 (50.5) | 712 (52.4) | 0.530 |
| Fever (>37.8°C) | 0 | 832 (46.6) | 202 (48.3) | 626 (46.1) | 0.450 |
| Dyspnea | 0 | 857 (48.0) | 238 (56.9) | 617 (45.4) | <0.001 |
| Typical symptoms |  |  |  |  |  |
| Malaise/fatigue | 0 | 665 (37.2) | 155 (37.1) | 504 (37.1) | 1 |
| Diarrhea | 0 | 291 (16.3) | 60 (14.4) | 229 (16.9) | 0.257 |
| Chills | 0 | 201 (11.3) | 30 (7.2) | 171 (12.6) | 0.003 |
| Myalgia | 0 | 189 (10.6) | 25 (6.0) | 161 (11.8) | 0.001 |
| Nausea | 0 | 154 (8.6) | 19 (4.5) | 133 (9.8) | 0.001 |
| Sputum | 0 | 140 (7.8) | 34 (8.1) | 106 (7.8) | 0.906 |
| Vomiting | 0 | 127 (7.1) | 24 (5.7) | 102 (7.5) | 0.263 |
| Abdominal pain | 0 | 95 (5.3) | 19 (4.5) | 76 (5.6) | 0.479 |
| Headache | 0 | 110 (6.2) | 21 (5.0) | 88 (6.5) | 0.335 |
| Rhinorrhea | 0 | 124 (6.9) | 30 (7.2) | 94 (6.9) | 0.942 |
| Tachycardia (>90/min) | 0 | 82 (4.6) | 34 (8.1) | 48 (3.5) | <0.001 |
| Sore throat | 0 | 113 (6.3) | 23 (5.5) | 88 (6.5) | 0.546 |
| Chest pain | 0 | 92 (5.2) | 17 (4.1) | 75 (5.5) | 0.296 |
| Dizziness | 0 | 95 (5.3) | 16 (3.8) | 78 (5.7) | 0.161 |
| Conjunctivitis | 0 | 12 (0.7) | 3 (0.7) | 9 (0.7) | 1 |
| Atypical symptoms |  |  |  |  |  |
| Anorexia | 0 | 598 (33.5) | 137 (32.8) | 457 (33.6) | 0.792 |
| Weakness | 0 | 519 (29.1) | 128 (30.6) | 388 (28.6) | 0.451 |
| Delirium | 0 | 449 (25.1) | 153 (36.6) | 295 (21.7) | <0.001 |
| Falls | 0 | 265 (14.8) | 67 (16.0) | 197 (14.5) | 0.489 |
| Functional decline | 0 | 142 (8.0) | 42 (10.0) | 100 (7.4) | 0.095 |
| Incontinence | 0 | 141 (7.9) | 31 (7.4) | 110 (8.1) | 0.73 |
| Weight loss | 0 | 24 (1.3) | 8 (1.9) | 16 (1.2) | 0.369 |
| Other | 0 | 2 (0.1) | 0 (0.0) | 2 (0.1) | 1 |

\*As collected on chart. IQR = interquartile range; SD = standard deviation

### Clinical presentation

At least one of the classic symptoms (Table 1) of fever (n=832, 46.6%), cough (n=929, 52.0%) or dyspnea (n=857, 48.0%) occurred in 1412 patients (79.1%). One or more typical symptoms occurred in 1263 patients (70.7%). The most common typical symptoms were malaise/fatigue (n=665, 37.2%), diarrhea (n=291, 16.3%) and chills (n=201, 11.3%). Atypical symptoms as defined by geriatric syndromes occurred in 1187 (66.5%) patients. The most common geriatric syndromes were anorexia (n=598, 33.5%), weakness (n=519, 23.9%), and delirium (n=449, 25.1%). Falls (n=265, 14.8%), functional decline (n=142, 8.0%) and urinary incontinence (n=141, 7.9%) were less common presentations of COVID-19. Functional decline as atypical presentation indicates a decline in functional status due to COVID-19 infection and not a baseline reduction in functional status. Atypical symptoms often occurred in the presence of classic (n=993, 83.7%) or typical symptoms (n=886, 74.6%). Approximately half of the cohort (n=738, 43.3%) presented with all 3 classes of symptoms (Figure 1). Atypical symptoms in isolation were uncommon (n=106, 6.2%).

**Figure 1:**
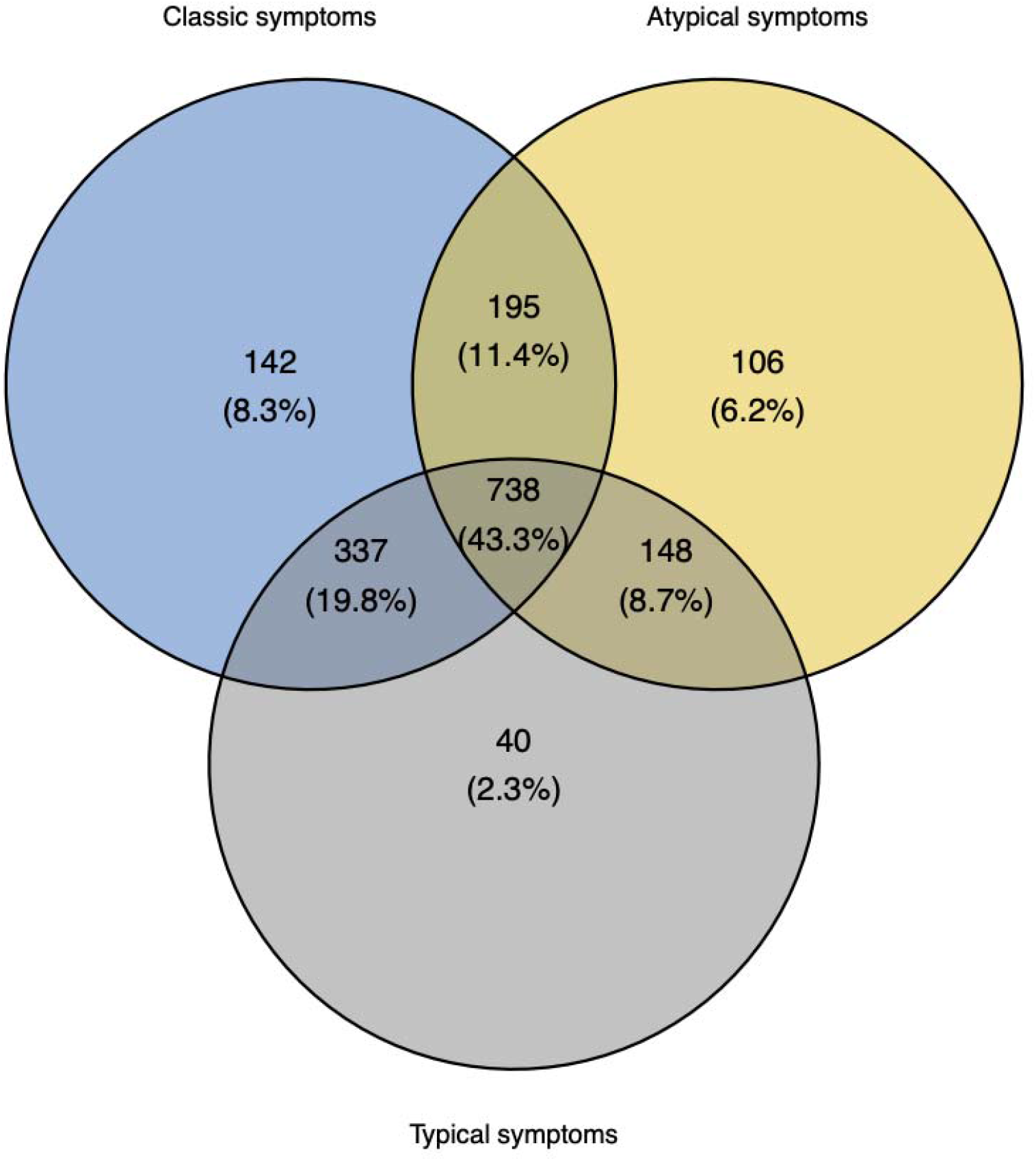
Distribution of patients presenting with classic, typical and atypical symptoms shown in a Venn diagram. Classic symptoms were defined by at least one of cough, fever or dyspnea. Typical symptoms included other symptoms such as rhinorrhea, chills, myalgias, diarrhea, etc. Atypical symptoms included geriatric syndromes such as delirium, falls, weakness, etc.

### Clinical presentation by pandemic wave

Hospitalized patients were younger (median age 75 years in wave 3 vs. 78.5 in wave 1, p<0.001) and less frail (mean CFS 4.09 in wave 3 vs. 5.21 in wave 1, p<0.001) in the later pandemic waves (Table 2). The prevalence of individual symptoms varied through the waves. For classic and typical symptoms, the prevalence of cough, abdominal pain, rhinorrhea, and tachycardia decreased through the waves, while the prevalence of dyspnea, chest pain, and dizziness increased. For atypical symptoms, the prevalence of anorexia, delirium, functional decline and incontinence decreased through the waves, but the prevalence of weakness increased.

**Table 2:**
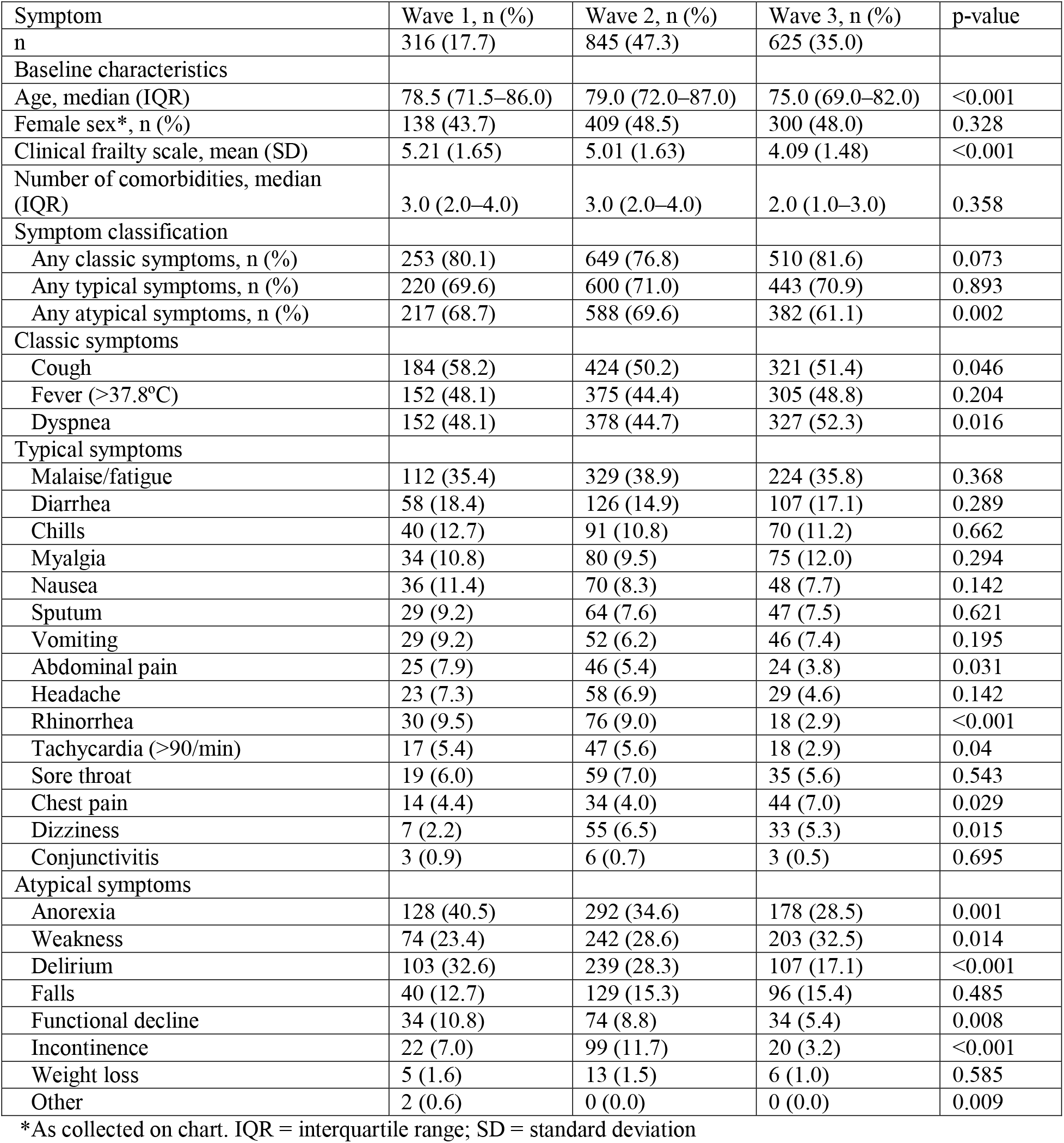
Prevalence of individual symptoms of wave of the pandemic.

### Clinical presentation and mortality

In-hospital death occurred in 418 patients (23.4%). Based on univariate analysis, patients who died (Table 1) were older (median age 82.0 years vs. 76.0, p <0.001), more likely to reside in LTC (21.2% vs. 12.5%, p=<0.001), and more frail (CFS mean 5.28 vs. 4.56, p<0.001). In univariate analysis, the presence of any classic (odds ratio [OR] 1.42, 95% confidence interval [CI] 1.07–1.90) or atypical symptoms (OR 1.28, 95% CI 1.01–1.63) was associated with in-hospital mortality. After adjusting for age, sex, CFS, and comorbidities (Table 3), only the presence of any classic symptoms was independently associated with in-hospital mortality (adjusted OR [aOR] 1.71, 95% CI 1.07–1.24).

**Table 3:**
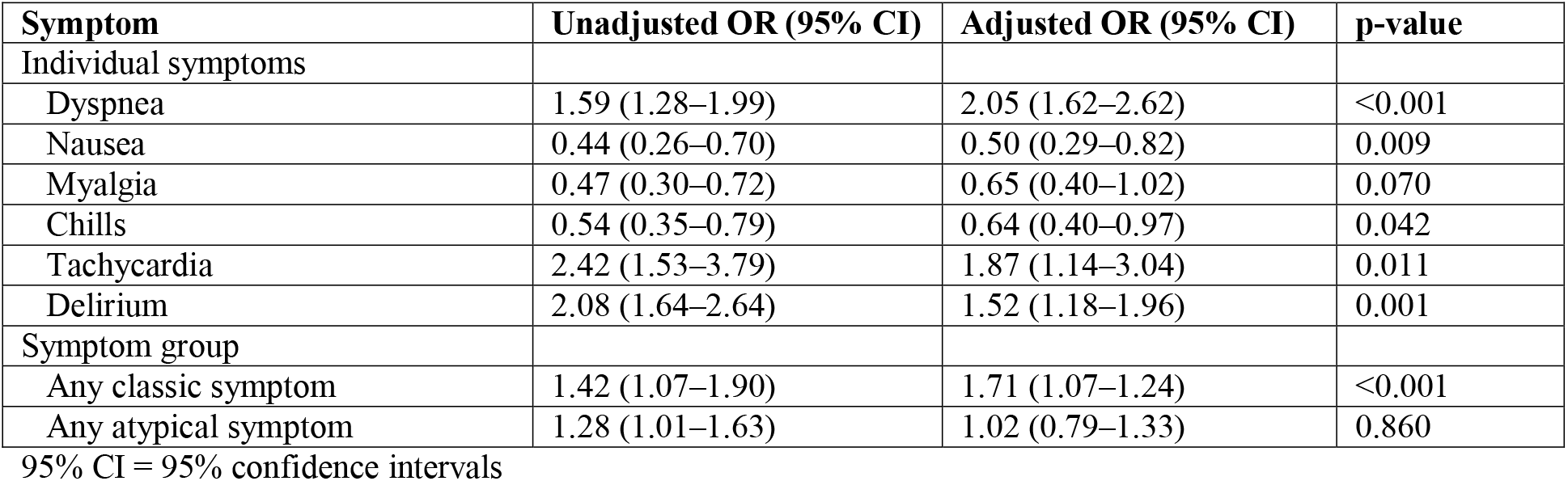
Unadjusted and adjusted odds ratios (OR) for symptoms that were significantly correlated with in-hospital mortality in the univariate analysis. Symptoms were adjusted for age (each 5 year increase), sex, clinical frailty scale, and total number of comorbidities using a multivariable model for each symptom.

For individual symptoms, the presence of dyspnea (aOR 2.05, 95% CI 1.62–2.62), tachycardia (aOR 1.87, 95% CI 1.14–3.04), and delirium (aOR 1.52, 95% CI 1.18–1.96) were independently associated with in-hospital mortality after adjusting for the same variables. The presence of nausea (aOR 0.50, 95% CI 0.29–0.82) and chills (aOR 0.64, 95% CI 0.40–0.97) was independently associated with lower mortality.

### Association of typical and atypical symptoms by age, frailty, place of residence and comorbidities

We combined classic and typical symptoms into a single category for this analysis, in order to plot it against atypical symptoms on a two-axis bubble plot. When grouped by age (Figure 2), those who were younger than 80 years had slightly more typical symptoms (45.6% vs. 42.8% in those aged ≥80 years), but the prevalence of atypical symptoms were similar. Frailty had a larger impact on atypical symptoms (Figure 2). Frail individuals (CFS >4) had more atypical symptoms compared to those who were not frail (61.1% vs. 38.9%). Looking at the place of residence in older adults prior to hospitalization (Figure 3), those from the community had fewer atypical symptoms (64.6% in community vs. 78.2% in LTC) compared to those from LTC. When analysed by individual comorbidity (Figure 4), those with a history of dementia and falls exhibited relatively more atypical symptoms (79.6% for both dementia and falls vs. 65.9% for other comorbidities) compared with other comorbidities. Those with a history of chronic obstructive pulmonary disease (COPD) had relatively more typical symptoms (91.2%) than atypical symptoms (56.6%). The comorbidities of hypertension, diabetes, cancer, chronic kidney disease and coronary artery disease clustered closely on the bubble plot, indicating a similar profile of typical and atypical symptoms.

**Figure 2:**
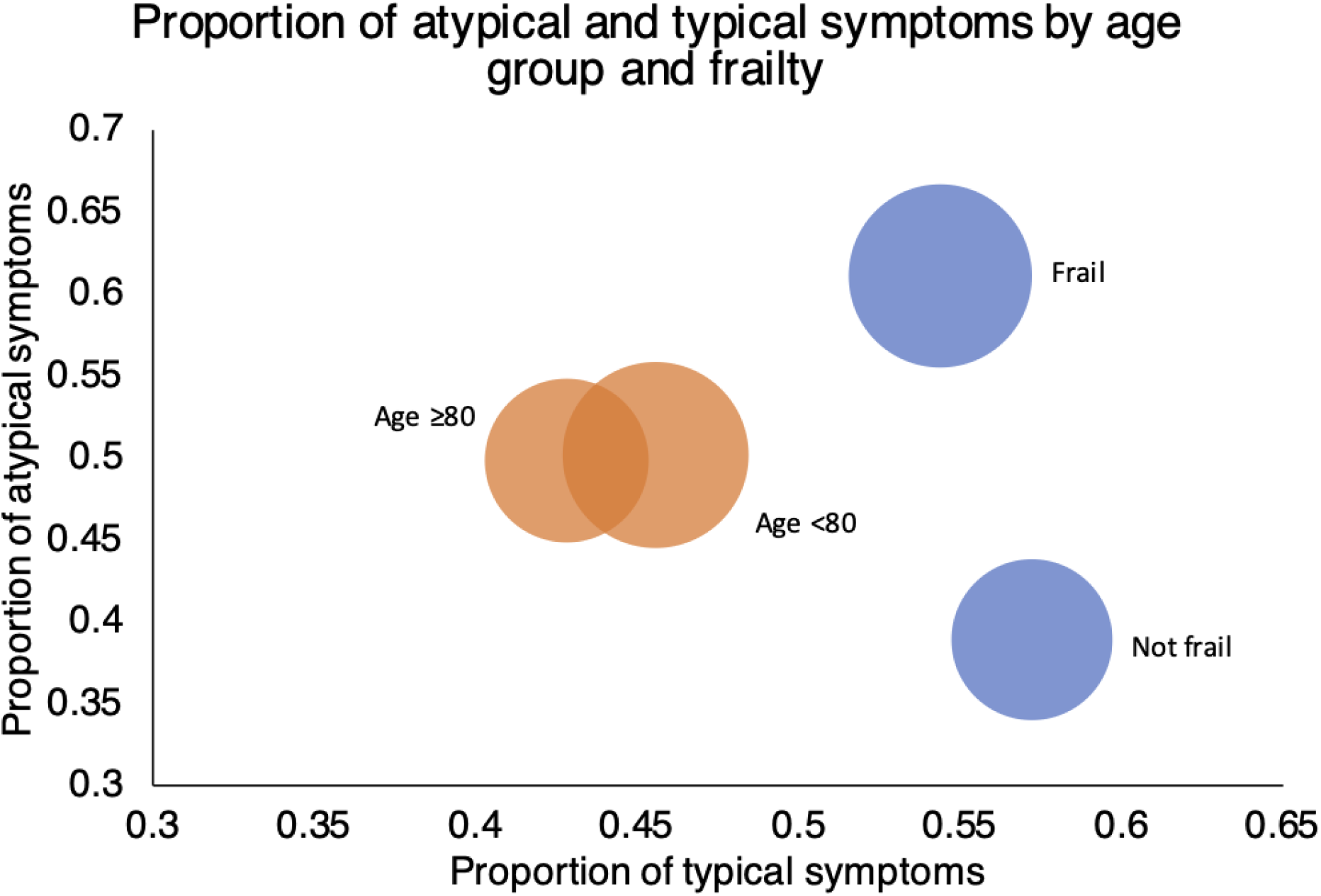
Bubble plot of proportion of atypical and typical symptoms by age group (<80 and ≥80 years) and frailty (clinical frailty scale ≤4 and >4 [frail]). The size of each circle corresponds to the sample size of each place of residence. Classic and typical symptoms were combined into a single category for this analysis. Atypical symptoms are defined by geriatric syndromes in our study (e.g. anorexia, delirium, weakness). Typical symptoms are non-geriatric syndromes (e.g. cough, dyspnea, fever).

**Figure 3:**
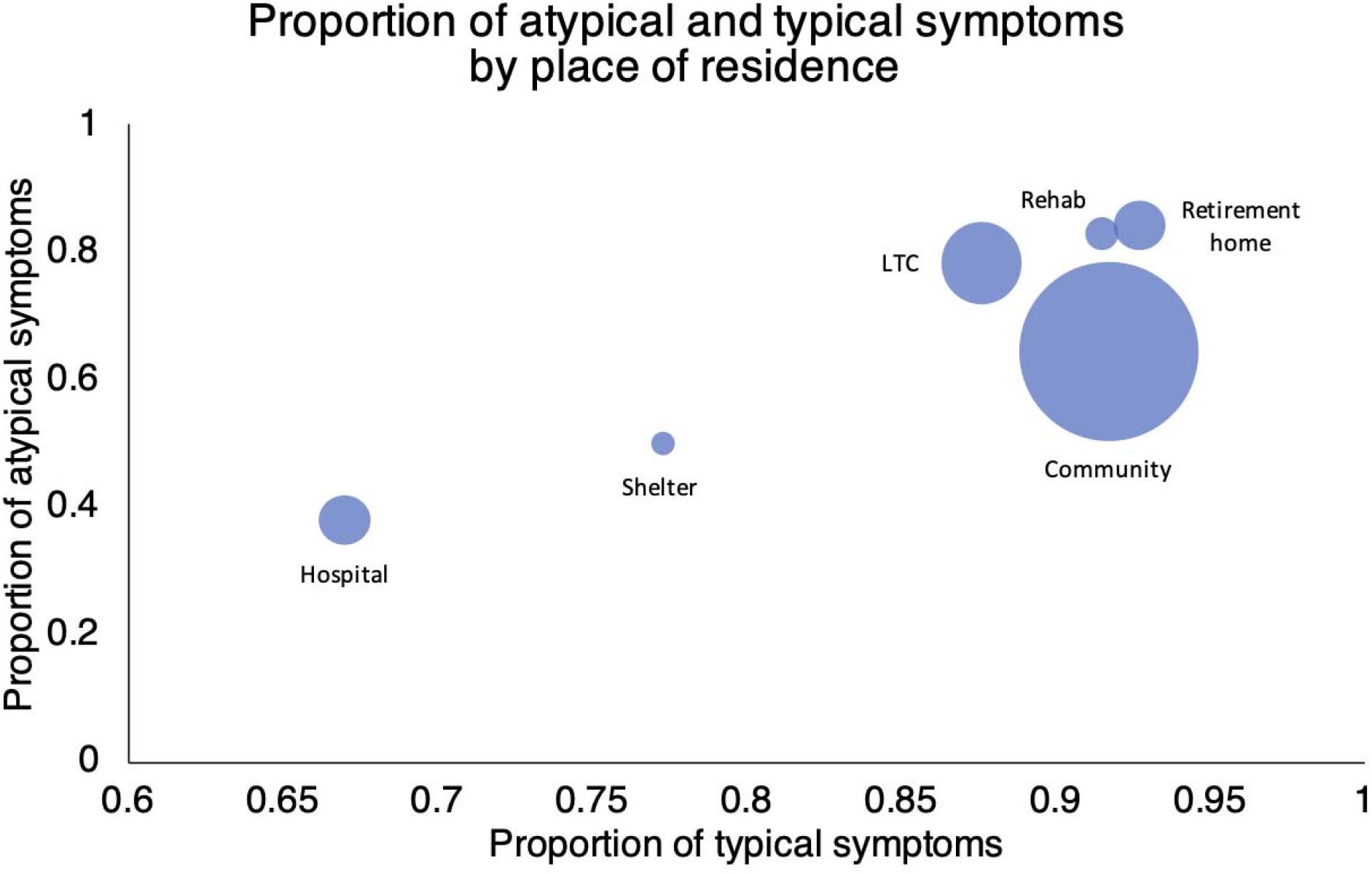
Bubble plot of the proportion of atypical and typical symptoms by place of residence (preadmission location) in older adults admitted to hospital with COVID-19. The size of each circle corresponds to the sample size of each place of residence. Classic and typical symptoms were combined into a single category for this analysis. Atypical symptoms are defined by geriatric syndromes in our study (e.g. anorexia, delirium, weakness). Typical symptoms are non-geriatric syndromes (e.g. cough, dyspnea, fever). LTC = long-term care.

**Figure 4:**
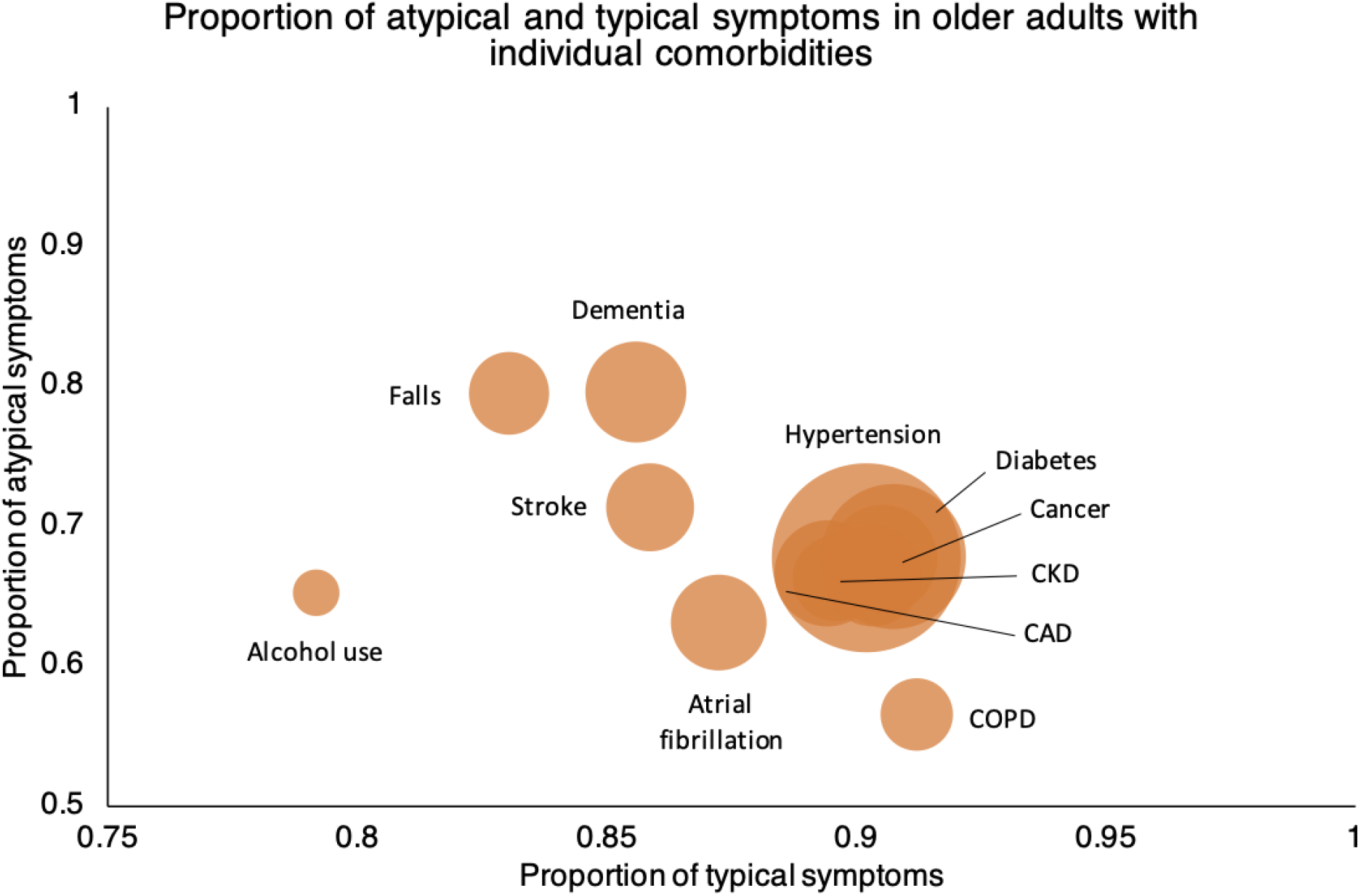
Bubble plot of atypical and typical symptoms by comorbidities. The size of each circle corresponds to the sample size of each place of residence. Classic and typical symptoms were combined into a single category for this analysis. Atypical symptoms are defined by geriatric syndromes in our study (e.g. anorexia, delirium, weakness). Typical symptoms are non-geriatric syndromes (e.g. cough, dyspnea, fever). CKD = chronic kidney disease; CAD = coronary artery disease; COPD = chronic pulmonary obstructive disease.

## Discussion

In this cohort study of consecutive older adults admitted to hospital and LTC with COVID-19, we found that atypical presentations were common, but most patients presented with an overlap of atypical, typical and classic symptoms. Given the inconsistency in defining atypical symptoms in the literature (Davis *et al*., 2020; Pritchard *et al*., 2020), we used three groups of symptoms to better describe the relationship between symptom types. The overlap in symptoms indicated that the presentation of COVID-19 in older adults was diverse. As such, a low threshold for testing is essential to detecting cases.

Fever (64–70%), dyspnea (71–73%) and cough (67–96%) were common symptoms in older adults (aged ≥65) admitted to hospital with community acquired pneumonia or influenza in pre-COVID studies (Riquelme *et al*., 1997; Walsh, Cox and Falsey, 2002). Comparatively, the prevalence of these symptoms in our cohort was lower (46.6–52.0%), suggesting that older adults with COVID-19 did not present as classically as respiratory infections of the past. A meta-analysis of COVID-19 studies in older adults from December 2019 to May 2020 revealed a weighted prevalence of 83% for fever, 60% for cough, and 42% for dyspnea. Increased virulence of the beta (B.1.351) and gamma (P.1) strains (Health Ontario, 2021), which predominated in the later waves in 2021, may have contributed to the higher proportion of dyspnea in our cohort (48.0%). Another explanation for higher dyspnea prevalence was the selection of sicker patients to be transferred from LTC to hospital given resource limitations at the time (CBC News, 2021) and from the community given increased recognition of symptoms requiring hospital treatment.

We defined atypical presentation as geriatric syndromes aligned with the clinical literature. Geriatric syndromes are phenotypes that arise from the interaction of frailty, multiple shared risk factors (e.g. cognitive impairment, gait instability, polypharmacy etc.) and the acute disease process (Inouye *et al*., 2007). They may not have a single direct pathophysiologic link to COVID-19, but rather the combination of infectious process on a susceptible older adult. Anorexia (44%) and delirium (44%) were the most common atypical symptoms in older adults hospitalized for community-acquired pneumonia in a study from Barcelona, Spain (Riquelme *et al*., 1997). In a cohort from Korea (Jung *et al*., 2017), weakness (27.3%) and delirium (25.4%) were most common in older adults hospitalized with pneumonia. These findings mirror the most common atypical symptoms in our cohort. Studies of atypical presentations in older adults with COVID-19 mostly report on delirium (Pranata *et al*., no date) and falls (Gawronska and Lorkowski, 2021), with fewer studies looking at anorexia, weakness and other geriatric syndromes. One problem is the lack of consensus definition on which geriatric syndromes are most important to capture (Inouye *et al*., 2007). Despite early concerns of isolated atypical presentations in older adults (Solanki, 2020), our study found few patients who presented with a geriatric syndrome alone without other symptoms (6.2%). The advocacy around atypical presentations raised awareness for case detection in this population, which helped improve protocols for LTC locally. Our findings highlighted the importance of understanding geriatric syndromes as part of disease presentations.

Using multivariable models, we determined that dyspnea, tachycardia and delirium were independently associated with in-hospital mortality. Systematic review of pooled univariate analyses found similar associations with mortality for dyspnea (Shi *et al*., 2021), tachycardia (Shi *et al*., 2021) and delirium (Pranata *et al*., no date). Dyspnea and tachycardia are likely reflective of hypoxia, which is an important independent predictor for disease severity and mortality in COVID-19 (Gallo Marin *et al*., 2021). Delirium is an independent predictor of mortality across medical and surgical patients in studies prior to COVID-19 (Aung Thein *et al*., 2020). These symptoms, including delirium, can help clinicians decide which patients to send to hospital in repeated waves of the pandemic. LTC outbreaks are occurring again in Ontario as a result of the omicron variant (McKenzie-Sutter, 2021) and clinicians can use these symptoms along with oxygen saturation to determine which patients need medications or acute care treatment. Conversely, the presence of nausea, chills, and myalgias were independently associated with decreased in-hospital mortality. Previous studies have not found these associations (McKenzie-Sutter, 2021), but most studies were done when the wild type version of the virus was circulating. It is possible that certain variants trigger differing symptoms (Health Ontario, 2021). Nausea, chills, and myalgias are also not related to respiratory status or hemodynamic instability, perhaps reflecting milder disease.

We further analysed clinical presentation by place of residence and comorbidities. The proportion of atypical symptoms were higher for those from LTC and those who had falls or dementia on history. These populations were likely more frail, which predisposes them to geriatric syndromes (Inouye *et al*., 2007). Patients from the community and those with common comorbidities like hypertension, diabetes, and coronary artery disease had relatively more typical symptoms as they were likely less frail. This finding suggests that in frail individuals with pre-existing geriatric syndromes attention should be given to atypical presentations and management should include prevention of geriatric complications (e.g. delirium, falls, medication adverse effects). Our analysis by frailty status and age confirmed that frailty was a relatively greater contributor to atypical presentations compared with age (Figure 2). Another finding from the bubble plot was that older patients with COPD often present with typical symptoms such as dyspnea or cough. Patients with COPD have upregulation of the angiotensin-converting enzyme 2 (ACE-2) receptor in their airway epithelial cells, which is the primary target for spike protein attachment during a SARS-CoV-2 infection (Higham *et al*., 2020). This allows for deeper and more severe infections in patients with COPD, which may result in predominantly respiratory symptoms, even early in the disease course (Higham *et al*., 2020). The varying clinical presentations with different comorbidities should be explored in future studies to further aid detection and mechanistic understanding of the disease.

This study was limited by its retrospective design, which did not allow for prospective delirium assessment or collection of symptoms. Agreement was not assessed for the chart abstraction process, but each chart assessor was trained and supervised by a physician investigator through the study. Frailty data were based on the documented pre-admission baseline functional status, and data were missing for some patients because a functional inquiry was not always included in clinical notes. We imputed the CFS for patients admitted from retirement home and LTC in a conservative way, which may underestimate true frailty. The CFS data were also imputed at the chart abstraction stage. SARS-CoV-2 variant data was not available at all sites, so we did not report it in this study, which limited our ability to analyse symptoms by variant.

Strengths of this study included the use of a three-category classification of symptoms that allowed determination of the contribution of geriatric syndromes. Compared to other studies we included more geriatric-focused symptoms, such as incontinence, falls, and functional decline. We also included consecutively admitted older adults to eight hospitals and had a consistent chart review process at all sites, guided by a physician at each site. Delirium was abstracted using a validated chart review method (Saczynski *et al*., 2014).

## Conclusion

Atypical presentations of COVID-19 in isolation are uncommon in older adults who were hospitalized. Of all symptoms, dyspnea, tachycardia and delirium were independently associated with mortality. Symptoms also varied by risk factors of individual patients, particularly by frailty status. Future research should be directed at understanding how COVID-19 interacts with different comorbid diseases and patient characteristics.

## Data Availability

All data produced in the present study are available upon reasonable request to the authors

## Acknowledgements

We thank Dr. Camilla Wong for providing training for chart abstraction; Dr. Samir Sinha and Dr. Rajin Mehta for assistance with funding and site project support.

## Authors and contributors

Conceptualisation, data curation, investigation, formal analysis, visualisation, validation, writing – original draft, and writing – review & editing: All authors (EKCW, JW, HZ, AC, AWZ, JB, AV, AB, RN, KP, TI, SES, BL).

Funding acquisition, methodology, project administration, resources – EKCW, JW, AV, AB, TI, RN, KP, SES, BL

Supervision – SES and BL

Underlying data was verified by EKCW, JW, RN, KP, TI, SES and BL. All authors confirm that they had full access to all the data in the study and accept responsibility to submit for publication.

## Declaration of interests

The authors have no conflicts to declare.

## Funding

Academic Health Science Centre Alternate Funding Plans (AFP) Innovative Funds from Unity Health Toronto and Baycrest Health Sciences; Sinai Heath/University Health Network Healthy Ageing and Geriatrics Program and its Geriatrics Summer Scholars Program; Division of Geriatric Medicine and General Internal Medicine, Sunnybrook Health Sciences Centre. The sponsor has no role in this study’s design, method, subject recruitment, data collection, analysis and manuscript. SES is funded by a Tier 1 Canada Research Chair. EKW is funded by the Clinician Scientist Training Program at the University of Toronto and the Vanier Scholarship from the Canadian Institutes of Health Research.

## Research in context

### Evidence before this study

Systematic reviews have demonstrated the prevalence of typical and atypical symptoms of COVID-19 in older adults. Atypical symptoms can have various definitions in the literature, including geriatric syndromes and other symptoms such as nausea, headache, and myalgia. Certain symptoms were known to be associated with a higher risk of mortality.

### Added value of this study

Our study uses a cohort of consecutively admitted older adults with COVID-19 in seven hospitals to determine the prevalence of atypical presentations as defined by geriatric syndromes. We separated the geriatric syndromes from other symptoms and showed that the occurrence of these symptoms in isolation was uncommon. We further analysed the evolution of symptom prevalence over three waves of the pandemic. We also determined that frailty was a bigger contributor to atypical symptoms compared with older age. Specific comorbidities also predisposed to a higher risk of atypical symptoms.

### Implications of all the available evidence

Awareness of the underlying patient characteristics can help guide clinicians in assessing for COVID-19 risk. Future studies should be done to determine the impact of comorbidities and frailty on disease presentations.

## Data sharing

Deidentified study data is available upon request.

## Notes

### Competing Interest Statement

The authors have declared no competing interest.

### Clinical Protocols

https://osf.io/k4g7a

### Author Declarations

Research ethics approval was obtained through Clinical Trials Ontario (3186-OPIA-Apr/2020-38044).

